# Test, track, and trace: How is the NHSX Covid app performing in a hospital setting?

**DOI:** 10.1101/2020.06.01.20116590

**Authors:** Joshua Filer, Daniel Gheorghiu

## Abstract

**Objective:** To assess the uptake and use of the trial contact tracing app developed by NHSX by healthcare workers.

**Design:** Cross-sectional study using survey questionnaire.

**Setting:** Healthcare industry: St Mary’s Hospital, a small NHS district hospital on the Isle of Wight, United Kingdom.

**Participants:** NHS staff members employed by the Isle of Wight NHS Trust.

**Results:** Of 3100 eligible staff members, 462 (~15%) responded to the survey. Of the respondents, 90% were aged between 31 and 65, and half had direct patient contact through their job role. Almost three quarters (73%) used social media apps on their smartphones. 421 out of 460 respondents had no trouble downloading and installing the NHSX Covid app on their smartphones. 20% of respondents were left confused by instructions to turn off Bluetooth when wearing PPE. Only 35 people either had to report symptoms or received an alert of contact with a suspected covid case. Of these over 20% were not clear what to do in such a situation.

**Conclusions:** The trial app has been embraced and adopted well. Many have experienced no problems with it. However, some healthcare workers have been unable to download or install the app due to compatibility issues and some have been left confused by having to turn off Bluetooth whilst wearing PPE.

This raises questions as to the effectiveness of the app for its intended purpose in contact tracing efforts.

**Recommendations:** We recommend that the wording of alerts and guidance provided by the app be made clearer and more accessible. We also recommend developments to the app to facilitate use by healthcare workers in a clinical setting. We also propose that ‘app instructors’ be made available in hospitals to ensure that patients and staff can access help and advice on use of the app.

## Introduction

The current covid-19 pandemic is the largest outbreak of an infectious disease that the NHS has faced in its history. In the early stages of the pandemic it was proposed that technology in the form of a smartphone app that can log travel and contact events could help improve the reliability and speed of contact tracing, thus helping in the efforts to isolate potential cases and reduce the transmission of the virus among populations.(1–3) These types of applications have been found to be useful in previous outbreaks,(4, 5) and as a result many countries have started using technology including smartphone apps to help with contact tracing.(2, 6, 7) In a press release from the department of health and social care at the beginning of May the UK government announced plans to trial such a smartphone-based app as part of their test, track and trace strategy.(8)

The Isle of Wight was chosen as a trial population due to its geography with a sizeable population and a single NHS trust that covers all NHS services on the Island. The initial rollout of the NHSX developed COVID-19 app began on the 5^th^ May 2020.(8)

The aim of this study was to assess the uptake, use and difficulties encountered whilst using this app by NHS staff at St Mary’s Hospital on the Isle of Wight.

## Methods

### Setting

St Mary’s Hospital is a district hospital located on the Isle of Wight and is run by the Isle of Wight NHS Trust. The trust currently employs 3100 people in a variety of clinical and non-clinical roles.(9)

### Study design

This cross-sectional study made use of a questionnaire, which was developed with questions relating to the download and use of the NHS X Covid app (Appendix 1). This was developed using SmartSurvey™ software (www.smartsurvery.co.uk) and was disseminated to staff members via email and in the form of an e-bulletin link on the hospital intranet on 13^th^ May 2020. The deadline for completion was 21^st^ May 2020. All staff members of Isle of Wight NHS Trust were eligible to complete the survey.

### Analyses

Basic descriptive statistics were calculated for each of the survey questions and common themes were identified and coded from the respondents’ comments.

### Patient and public involvement

As the study was based on data from anonymised questionnaire responses of NHS trust staff members, patients, and members of the public were not involved.

## Results

Of the roughly 3100 people who work for the Isle of Wight NHS trust, 462 people (~15%) completed the survey. Respondents included a range of people of different ages, although 90% were between the age of 31 and 65 (Figure 1). There was a reasonable representation of all services across the trust, including roughly equal proportions of staff who either have direct or no patient contact (51% vs 49% respectively, Figure 2).

**Figure 1.**
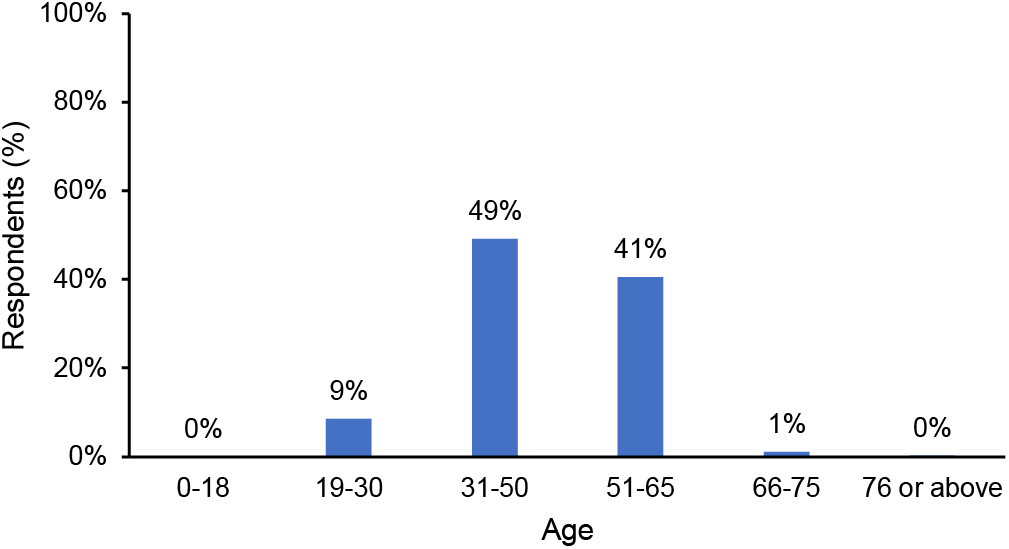
Age distribution of survey respondents.

**Figure 2.**
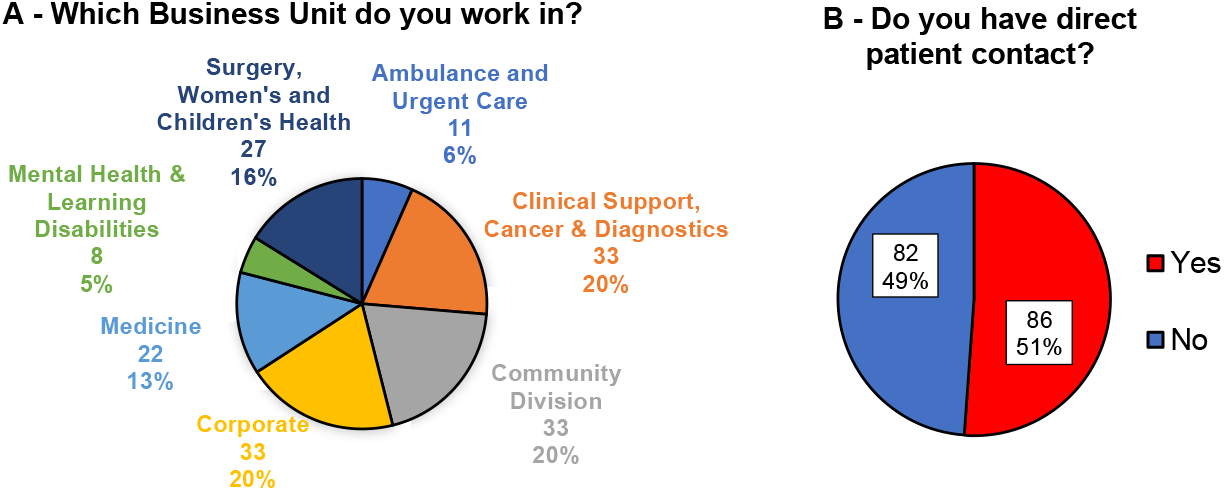
A – Range of different job roles of survey respondents within the Isle of Wight NHS trust (Respondents categorised by Business Unit worked for); and B – The proportion of respondents who have direct patient contact.

The majority of respondents had downloaded and installed the app on their smartphones (93%, 431 people). Of these, most had no problems locating, doing so (92%, 421 people). Small numbers of people had difficulties either downloading the correct app (4 people) or had smartphones which did not support use of the app (12 people) (Figure 3).

**Figure 3.**
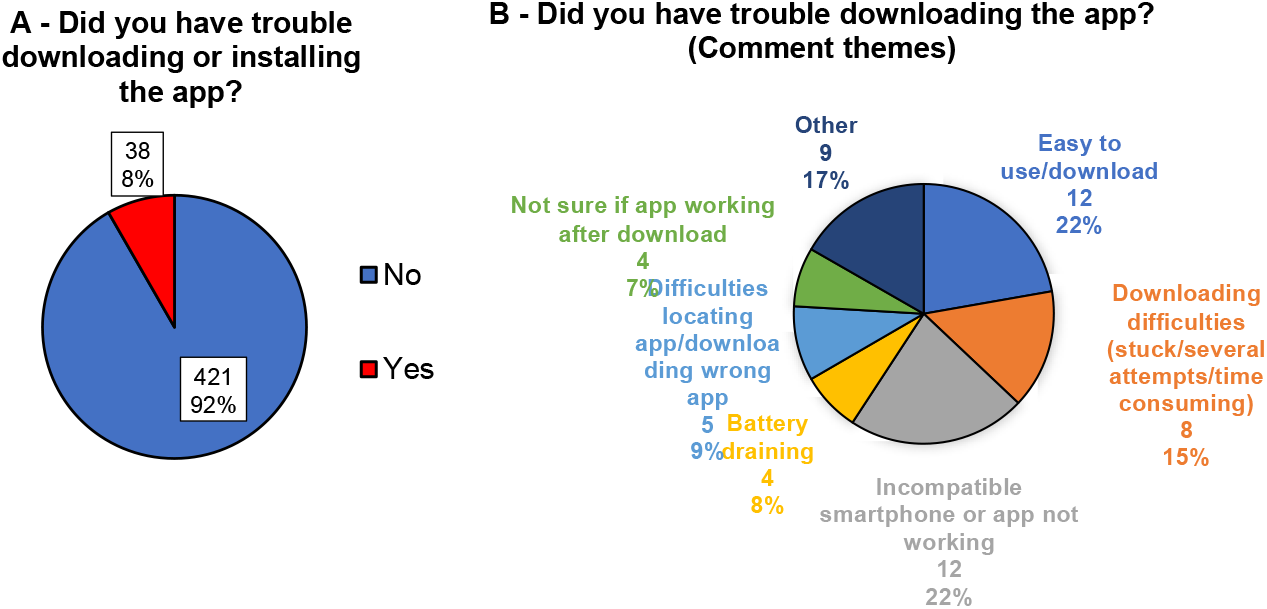
A – Proportion of respondents who had trouble downloading the app, B – Common themes reported by respondents when asked if they had trouble downloading the app (54 people provided comments).

Approximately three quarters (74%, 340 people) of respondents use other social media apps on their smartphones on a regular basis. A common theme in the comments of respondents was that they had problems using other apps including these social media platforms since downloading the NHSX Covid app, and that the battery life of their smartphones was impaired.

Approximately 20% of respondents were left confused about what level of personal protective equipment (PPE) use would require them to switch off their Bluetooth (Figure 4). People reported different interpretations of this instruction with some turning it off whilst wearing a mask of any kind, whilst others would only turn it off when wearing full PPE with FFP3 masks. Some staff members would turn off Bluetooth only whilst on wards with patients who had tested positive for covid-19, whereas others would turn it off when in work. We had reports that some users leave it switched on in locker rooms whilst at work and some turn it off whilst at home. Others forget to turn it off at all, even whilst wearing full PPE.

**Figure 4.**
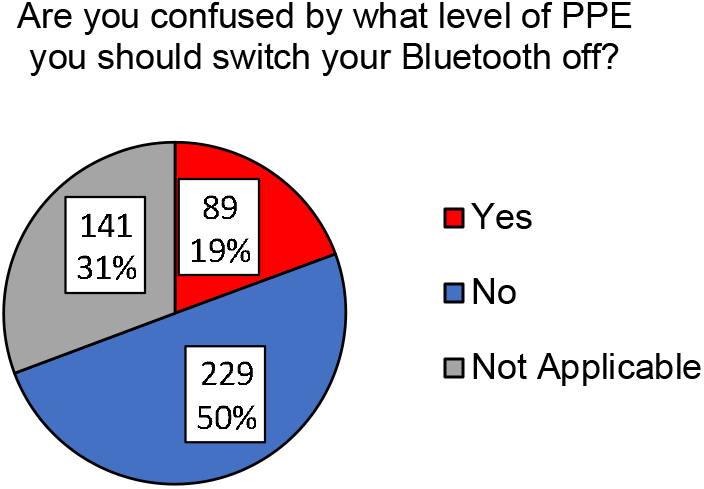
The proportion of respondents who were confused by the instruction to turn off their Bluetooth when wearing PPE. NA = Proportion of those who are not required to switch Bluetooth off or on during their working duties.

A small number of people (35, 8%) either had to report symptoms or received an alert that a contact had either reported symptoms or had tested positive for covid-19 (Figure 5). Over 20% of these stated that “when alerted by the app it was unclear what to do”. Many reported that there were not adequate options for reporting symptoms or test results. Many also reported that advice both on the app, the government website and that given out by the NHS trust was unclear or conflicting.

**Figure 5.**
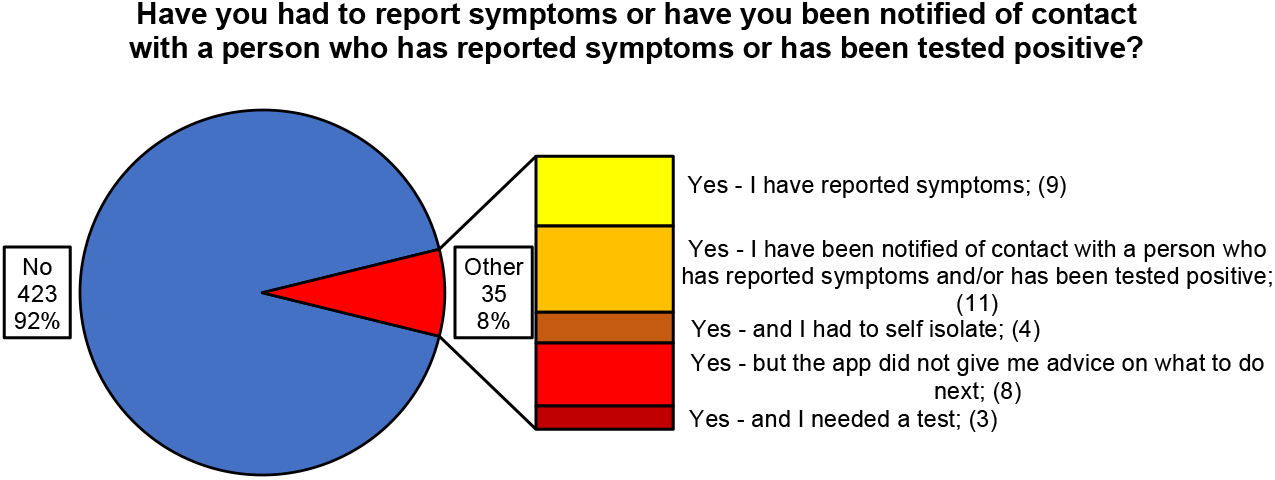
Proportion of respondents who have had to report symptoms or received an alert of contact with someone who had either symptoms or a positive covid-19 test.

## Discussion

The current Covid-19 pandemic is the largest outbreak of an infectious disease that the NHS has faced in its history. At time of writing, the acute respiratory illness caused by the novel coronavirus (SARS-CoV-2) has spread from China to over 180 countries, with over 4.3 million confirmed cases and over 290,000 deaths globally.(10, 11) In the UK there have been over 230,000 confirmed cases and 33,263 deaths.(11, 12)

Many countries have been in prolonged lockdown with people’s movements and activities restricted in efforts to slow the transmission of the virus and limit the number of deaths. This has had an enormous impact upon the global economy with many people losing their jobs and significant government bailouts being introduced. In order to come out of lockdown and restart the economy safely whilst limiting the spread of this highly contagious respiratory disease, it is widely believed that contact tracing and targeted isolation is needed.(1, 6, 13, 14) In the healthcare setting where exposed but asymptomatic healthcare workers can unwittingly spread the virus to their patients, contact tracing is perhaps even more important to help limit the outbreak.(15) However, contact tracing is very labour intensive and is reliant upon the people who have tested positive remembering everyone who they have had contact with.(1, 6, 14) This is a difficult task and is made more difficult when the asymptomatic incubation period is up to around 14 days from initial exposure.(1, 14) The initial rollout of the NHSX developed COVID-19 app began on the 5^th^ May 2020.(8) Initial concerns were that the Isle of Wight population, due to its higher percentage of over 65 year olds, might not achieve the recommended download threshold of 60% which could bring an outbreak under control.(16)

Although the population of the Isle of Wight is approximately 141,000,(17) when estimates of those who own a smartphone and are of legal age to download the app are considered, it has been estimated that around 80,000 would be theoretically able to download and use the app.(18)

On 11^th^ May 2020 it was announced that the app had been downloaded approximately 55,000 times. This rose to around 70,000 downloads by 18^th^ May and is thought to represent approximately 65% of those on the island who could download the app. This uptake almost reaches the 60% of population threshold mark within a week of the app’s release. This encouraging uptake came despite early reports that the government’s anticipated coronavirus tracing app had failed crucial security tests and was not yet safe enough to be rolled out across the UK.(18–20) Although application data security concerns were certainly a commonly reported theme in our survey, this did not seem to effect uptake of the app.

Possibly the biggest failure of the app for healthcare workers is the confusion about when to stop track and tracing, whilst wearing PPE and the wording around PPE used by the app. Our trust guidelines concerning PPE follow the guidance provided by Public Health England.(21) When starting the app, the user is given important instructions for healthcare workers: “Your interaction at work should not be captured when you are wearing personal protective equipment.” Using the information button integrated in the app takes the user to the official website (www.covid19.nhs.uk). Under the subheading “Common questions about the app”, the user can access additional advice to the question; “Should healthcare workers use the app.” The answer here states: “It is important for health and care workers to install the app at all times except when they are wearing appropriate PPE and come in close proximity with a patient with confirmed or suspected covid-19. When wearing PPE for confirmed or suspected Covid-19 patients, staff should turn their Bluetooth off.(22) The multiple stages required to get clarification, and the wording of the answers provided is not user friendly. This likely contributes to the confusion reported by almost 20% of respondents regarding the circumstances that require healthcare workers to switch off Bluetooth.

As a result of this confusion and the inconvenience of switching on and off Bluetooth when moving between different clinical situations, there have been comments that lead us to believe that not all users are correctly using the app. When the app is used incorrectly, for example when the user fails to correctly activate or deactivate Bluetooth, or leaves their phone in locker rooms or offices, there is a real risk that there will be false notifications of contact. Additionally, incorrect use of the app may lead to failures to capture potential contacts thus rendering the app inaccurate in contact tracing. Furthermore, the fact that end users cannot trace potential contacts (as location data of the possible contact and details of the contact is not provided by an alert) valid risk assessment of real contact is difficult, resulting in difficult decision-making in relation to the actions that are required (i.e. to self-isolate, or to arrange for asymptomatic testing).

The minimum recommended PPE for primary, outpatient, community and social care by setting for the NHS and independent sector consists of a fluid resistant mask.(21) Although our survey questions cannot give any correlation to the type of PPE respondents were wearing and their reported problem with the app, the comments given in our survey indicate that at least some of the respondents would fall in the above category of recommended PPE, and were also part of the 20% who were confused about when to switch Bluetooth off. One could argue that some members of the general public are wearing a similar level of PPE as healthcare workers (i.e. fluid resistant masks) when out of the house for exercise or other essential activities. Often these people may not be able to maintain the 2-meter social distancing. Therefore, one could argue that those individuals of the general public wearing PPE, should also switch Bluetooth off in certain circumstances. If this isn’t necessary then why are healthcare workers being asked to switch Bluetooth off when wearing the above-mentioned PPE?

Over 20% of the respondents who were notified of a contact reported that the app did not give them advice on what to do next. This indicates that the advice function was not operating. Being unclear what to do when you receive an alert of a possible contact is clearly unhelpful and renders the app ineffective in limiting the spread of disease.

The following comment from a respondent of our survey possibly sums up the performance of the app in the healthcare sector the best:

“Following the trial, all we know is that the app works on certain people’s phones in terms of sending messages but we will not be able to derive any data regarding whether contact tracing reduces the number of new cases. Also, the app will require certain alterations to be beneficial and accurate especially in the healthcare sector.”

## Recommendations

1. We would recommend the provision of unified wording and clear recommendations for healthcare workers using PPE.
2. We would recommend an adaptation to the app specifically for the healthcare sector, allowing a more informed risk assessment for the end user.
3. In order to provide clear and standardised advice, we have introduced an “App instructor” at the reception desk of the Hospital who will greet patients and staff, asking them if they need any help downloading or working the app.

## Data Availability

The data used in this study will be made available upon request

## Key Messages

### What is already known on this topic

The new app is currently being trialled on the Isle of Wight so little is known about its use and problems. The NHSX Covid app is being developed and tested for use in helping contact tracing during the coid-19 virus pandemic.

### What this study adds

This study highlights the problems that have been encountered by healthcare workers whilst using the app in the course of their clinical duites. These include difficulties downloading and installing the app, confusion on when to turn off Bluetooth whilst wearing PPE and lack of clear advice on what to do when notified of contact by the app.

## Acknowledgements

We would like to thank the Communications department of the Isle of Wight NHS Trust for their help and support in disseminating the survey questionnaire.

## Appendix 1

Survey questions disseminated to staff at the Isle of Wight NHS trust 1 Have you downloaded the app?

|  |  |  |
| --- | --- | --- |
| 1 | Have you downloaded the app? |  |
|  | 1 | Yes - and I am using it |
|  | 2 | Yes - but did not feel well informed about it |
|  | 3 | Yes - but I have deleted it again |
|  | 4 | No - I don't want to for any reason |
|  | 5 | No - I don't have a smartphone or any device compatible for the app |
|  | 6 | No - I don't trust the security |
|  | 7 | No - I don't think the app will make any difference |
| 2 | What is your age? |  |
|  | 1 | 0-18 |
|  | 2 | 19-30 |
|  | 3 | 31-50 |
|  | 4 | 51-65 |
|  | 5 | 66-75 |
|  | 6 | 76 or above |
| 3 | Which Business Unit do you work in? |  |
|  | 1 | Ambulance and Urgent Care |
|  | 2 | Clinical Support, Cancer & Diagnostics |
|  | 3 | Community Division |
|  | 4 | Corporate |
|  | 5 | Medicine |
|  | 6 | Mental Health & Learning Disabilities |
|  | 7 | Surgery, Women's and Children's Health |
| 4 | Do you have direct patient contact? |  |
|  | 1 | Yes |
|  | 2 | No |
| 5 | Did you have any trouble to download or install the app? |  |
|  | 1 | No |
|  | 2 | Yes - I downloaded the wrong app by mistake |
|  | 3 | Yes - The app does not work on my phone or is not supported by its software |
|  | 4 | Other |
|  |  | Comments: |
| 6 | Are you confused by what level of PPE you should switch your Bluetooth off? |  |
|  | 1 | Yes |
|  | 2 | No |
|  | 3 | Not Applicable |
| 7 | If applicable, when do you switch it off? |  |
| 8 | Anything else you can tell us about the app? |  |
| 9 | Do you use any other social media apps on your phone? (i.e. Facebook, Twitter, Instagram, Strava etc) |  |
|  | 1 | Always |
|  | 2 | Never |
|  | 3 | Sometimes |
| 10 |  | Have you had to report symptoms or have you been notified of contact with a person who has reported symptoms or has been tested positive? |
|  | 1 | Yes - I have reported symptoms |
|  | 2 | Yes - I have been notified of contact with a person who has reported symptoms and/or has been tested positive |
|  | 3 | Yes - and I had to self-isolate |
|  | 4 | Yes - but the app did not give me advice on what to do next |
|  | 5 | Yes - and I needed a test |
|  | 6 | No |

